# Early Prediction of COVID-19 Severity Using Extracellular Vesicles and Extracellular RNAs

**DOI:** 10.1101/2020.10.14.20212340

**Authors:** Yu Fujita, Tokio Hoshina, Juntaro Matsuzaki, Tsukasa Kadota, Shota Fujimoto, Hironori Kawamoto, Naoaki Watanabe, Kenji Sawaki, Yohei Sakamoto, Makiko Miyajima, Kwangyole Lee, Kazuhiko Nakaharai, Tetsuya Horino, Ryo Nakagawa, Jun Araya, Mitsuru Miyato, Masaki Yoshida, Kazuyoshi Kuwano, Takahiro Ochiya

## Abstract

The clinical manifestations of COVID-19 vary broadly, ranging from asymptomatic infection to acute respiratory failure and death. But the predictive biomarkers for characterizing the variability are still lacking. Since emerging evidence indicates that extracellular vesicles (EVs) and extracellular RNAs (exRNAs) are functionally involved in a number of pathological processes, we hypothesize that these extracellular components may be key determinants and/or predictors of COVID-19 severity. To test our hypothesis, we collected serum samples from 31 patients with mild COVID-19 symptoms at the time of their admission. After standard therapy without corticosteroids, 9 of the 31 patients developed severe COVID-19 symptoms. We analyzed EV protein and exRNA profiles to look for correlations between these profiles and COVID-19 severity. Strikingly, we identified three distinct groups of markers (antiviral response-related EV proteins, coagulation-related markers, and liver damage-related exRNAs) with the potential to serve as early predictive biomarkers for COVID-19 severity. Among these markers, EV COPB2 has the best predictive value for severe deterioration of COVID-19 patients in this cohort. This type of information concerning functional extracellular component profiles could have great value for patient stratification and for making early clinical decisions about strategies for COVID-19 therapy.

## Introduction

Severe acute respiratory syndrome coronavirus 2 (SARS-CoV-2) is responsible for a rapidly-unfolding pandemic that is overwhelming health care systems worldwide ^1^. The majority of Coronavirus-disease 2019 (COVID-19) patients exhibit mild clinical symptoms, including fever, cough, and sputum. In some cases, more severe symptoms such as dyspnea and/or hypoxemia occur after 1 week, with 50% of these severely-affected patients quickly progressing to systemic, life-threatening disorders, including acute respiratory distress syndrome (ARDS), septic shock, refractory metabolic acidosis, coagulation disorders, and multiple organ failure ^2^. These problems lead to both mild and severe respiratory manifestations, the latter being prominent in the elderly and in those with underlying medical conditions such as cardiovascular and chronic respiratory disease, diabetes, and cancer ^2^. The main clinical feature of severe COVID-19 is the onset of ARDS ^3^. It has been reported that the rate of occurrence of ARDS in patients with severe COVID-19 is 15.9%-29% ^2,4^. The immune response that is vital for the control and resolution of SARS-CoV-2 infections can also lead to cellular damage in association with systemic deteriorations. Recent studies suggest that an exaggerated inflammatory response known as cytokine storm may play a key role in this process ^5^. The variability of patient responses to SARS-CoV-2 infection makes it difficult to identify individuals who may be most at-risk for severe responses to the disease and to create early strategies for treating the disease. Although PCR-based detection of SARS-CoV-2 is useful for accurately diagnosing the disease, there is no available tool for effectively predicting disease severity or likely trajectories of illness progression ^6^.

Early identification of COVID-19 patients who may experience severe disease deterioration is of great significance for improving clinical approaches that will result in reduced mortality. Accordingly, the purpose of our study has been to identify extracellular components that can serve as useful biomarkers for predicting disease progression in COVID-19 patients. Toward this end we have focused on identifying the profiles of extracellular vesicle (EV) proteins and extracellular RNAs (exRNAs) in serum samples taken from COVID-19 patients with mild symptoms at the time of their admission for medical treatment.

Extracellular vesicles (EVs), including exosomes and microvesicles, contain numerous and diverse bio-molecules such as RNAs and proteins within the vesicle’s lipid bilayer ^7^. This stable bilayer construction enables intact EVs to circulate through body fluids. Thus, EVs can transfer their cargo to target cells as a means of regulating target cell participation in both physiological and pathological processes, including host immune responses. EVs are characterized by a number of exosomal markers, including members of the tetraspanin family (CD9, CD63, CD81), components of the endosomal sorting complex required for transport (ESCRT) (TSG101, Alix), heat shock proteins (Hsp60, Hsp70, Hsp90), and RAB proteins (RAB27a/b)^8^. Accumulating evidence suggests that the size, membrane composition, and contents of EVs are highly heterogeneous, depending on the cellular source. Furthermore, EV cargoes can be dynamically altered by microenvironmental stimuli including viral infection. The EVs produced by both immune and non-immune cells can be responsible for regulating the nature of the immune response as it relates to inflammation, autoimmunity, and infectious disease pathology ^9^. EVs derived from virus-infected cells have been shown not only to modulate immune cell responses, but also to spread the viral infection via delivery of the viral genome and protein particles to healthy cells ^10^. Intriguingly, EVs have receptors for coronavirus entry such as CD9 ^11^ and angiotensin-converting enzyme 2 (ACE2) ^12^, suggesting that EVs may also have a role in modulating or mediating SARS-CoV-2 infection. For these reasons, evaluating the molecular profiles of circulating EVs in COVID-19 patients may provide clues for understanding the host antiviral immune response and the mechanisms that determine disease trajectory and patient deterioration.

Studies of extracellular RNA (exRNA) have recently broadened into an important area of research with relevance to disease biomarker discovery and therapeutics ^13^. Most exRNAs are protected from degradation in biofluids via incorporation into EVs or into complexes with lipids and proteins. The variety of exRNA species that have been identified include messenger RNAs (mRNAs) and non-coding RNAs such as microRNAs (miRNAs), small nuclear RNAs, transfer RNAs, and long non-coding RNAs (lncRNAs). During the early stages of viral infection, host non-coding RNAs such as miRNAs are produced and released as a part of the antiviral response that is aimed either directly or indirectly at targeting viral transcription, translation, and replication ^14^. ExRNAs mediate a complex network of interactions between the virus and infected host cells, representing a pivotal role in modulating the host innate immune system ^15^. These findings suggest that exRNAs may have the potential to serve as biomarkers for evaluating the antiviral responses mounted by COVID-19 patients.

## Results

### Baseline clinical characteristics of the patient cohort

For this retrospective, single-center study, 31 SARS-CoV-2 infected patients and 10 uninfected healthy donors were included in the patient cohort. At the time of patient admission for treatment, serum samples were taken from all 41 individuals as a source of material for identifying biomarkers (**Table S1**). The severity of COVID-19 disease in each patient was graded using the WHO ordinal outcome scale of clinical improvement (**Table S2**). At the time of admission, all COVID-19 patients were scored as having a mild status (WHO score = 3). Based on the clinical course of disease progression after admission, we divided the 31 COVID-19 patients into two groups: Group 1 included patients who retained their mild status (WHO score ≦ 4), and Group 2 included patients who progressed to severe status (WHO score ≧ 5) (**Figure S1a**). All patients in Group 1 were subsequently discharged in good health from the hospital or transferred to the local medical facility for further observation. In Group 2, two patients (22.2%) died from COVID-19 complications (pulmonary embolism (*n*=1) or ARDS (*n*=1)), and two patients (22.2%) remained hospitalized at the time of writing. All other patients in Group 2 were discharged in good health from the hospital following treatment (**Table S1**). Clinical parameters were comparable among the groups (**Table S3**). Healthy donors and COVID-19 patients (Group 1 & 2) did not differ with respect to age, sex, body mass index (BMI), smoking index, levels of blood urea nitrogen (BUN), creatinine (Cr), and alanine aminotransferase (ALT), history of hypertension, diabetes mellitus, dyslipidemia, and coronary heart disease (*P*>0.05). Significant differences were observed in white blood cell (WBC) count and levels of C-reactive protein (CRP) (*P* <0.05). Comparison of COVID-19 patients in Group 1 and Group 2 revealed no significant difference in sex, BMI, WBC count, BUN, Cr, creatine kinase (CK), D-dimer, and fibrinogen, history of hypertension, diabetes mellitus, dyslipidemia, and coronary heart disease (*P* >0.05). However, significant differences were observed with respect to age, smoking index, and levels of CRP and ALT (*P* <0.05). We selected these 4 parameters for Pearson’s correlation analysis to identify correlations between the three groups in terms of age, smoking index, CRP, and ALT (**Figure S1b**). There were significant correlations in age, CRP, and ALT between the three groups (*P* trend <0.05).

### Predictive value of 4 clinical parameters for COVID-19 severity

We performed receiver operating characteristic (ROC) analysis on combined Group1 and Group 2 to provide a robust test of sensitivity, specificity, and area under the curve (AUC) for our set of four predictive parameters (**Figure S1c**). The AUC values for the four factors (age, smoking index, CRP, and ALT) were 0.90 (95% CI: 0.79-1.00), 0.69 (95% CI: 0.48-0.89), 0.77 (95% CI: 0.60-0.94), and 0.72 (95% CI: 0.51-0.94), respectively. These findings suggest that at the time of admission to the hospital, age and CRP, having AUC values > 0.75, can provide the greatest discrimination between patients who will develop mild versus severe COVID-19.

Based on the ROC analysis, optimal cutoffs for the 4 parameters were identified via use of the Youden index (**Table S6**). For further analysis, we used these cutoffs to separate COVID-19 patients into low and high groups for comparing the incidence of severe COVID-19 related events. Kaplan-Meier curves were constructed for each of the four parameters to compare the high and low groups for time-to-onset of a severe event (beginning with the day of admission) (**Figure S1d**). The progression-free times for age and CRP were significantly shorter in the high group than in the low group (*P*=3.9×10^−6^; *P*=0.014, respectively). These findings suggest that the parameters of patient age and CRP can be useful at the time of admission to predict the incidence of future severe COVID-19 related events. Currently, age and/or CRP levels are consistently used in the clinic for risk stratification to predict the potential severity of COVID-19 progression ^2,16^. Results with our cohort of 31 COVID-19 patients are therefore representative of the overall status of patients infected with SARS-CoV-2.

### LC-MS analysis of proteome profiles from EVs in serum samples from COVID-19 patients and uninfected controls

In clinical settings, analysis of EVs from liquid biopsies has gained attention as a potential means of identifying diagnostic and prognostic biomarkers for various diseases. However, this strategy has not yet been widely used due to a lack of standardized methods for isolating EVs from patients. To improve on this situation, we have employed an immunoprecipitation (IP) based method that targets surface marker proteins for rapid and specific isolation of EVs. The use of IP in the presence of a chelating reagent improves the yield and purity of CD9^+^ or CD63^+^ positive EVs from serum samples. The resulting EV preparations are suitable for subsequent EV proteome analysis by LC-MS (**Figure 1a**). From our total of 41 serum samples, LC-MS analysis identified 1676 proteins, following exclusion of proteins that were not present in all samples. Of these 1676 proteins, a total 723 proteins were present at different levels between the three groups (*P*<0.05; one-way ANOVA). To compare the pattern of expression of these 723 EV proteins among the three patient cohorts, we used unsupervised multivariate statistics based on principal component analysis (PCA) mapping. PCA plots the first and second principal components (PC1 and PC2) using all the term frequency from the LC-MS data, showing a certain degree of separated trends between the three groups (**Figure 1b**). This differentiation could be described by the first PC1, which accounted for 28.1% of the variance. The second PC2 accounted for 14.5% of the variance. This observation indicated that the EV proteome profiles of serum in COVID-19 patients deviated considerably from those uninfected donors. Although a little overlap or dispersity was demonstrated, we found obvious EV proteomic differences between Group 1 and Group 2 in the PCA scores plot.

**Figure 1.**
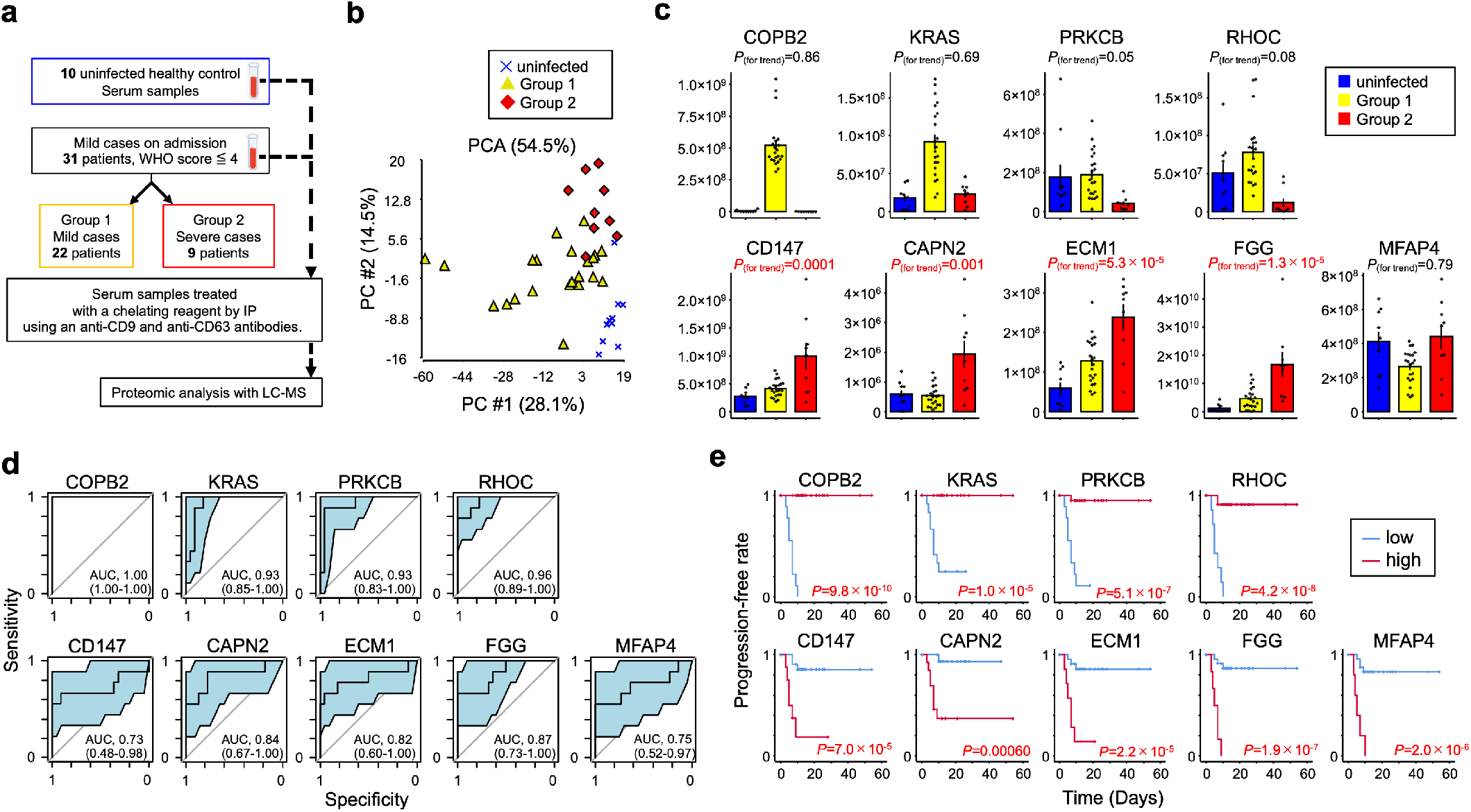
EV proteomes for early prediction of COVID-19 severity. (**a**) Work flow for LC-MS identification of proteomes from CD9+/CD63+ EVs obtained from serum samples of 31 mild COVID-19 subjects (Group 1: *n*=22, Group2: *n*=9) and 10 uninfected healthy controls. (**b**) PCA map of 723 proteins from the three subject groups. (**c**) Correlations of COPB2, KRAS, PRKCM, RHOC, CD147, CAPN2, ECM1, FGG, and MFAP4 between the three subject groups. *P* values for trend by *Pearson’s* correlation analysis. Error bars represent mean SEM. (**d**) AUC values (95% CI) for 9 EV proteins evaluated by ROC analysis. (**e**) Kaplan-Meier curves for 9 EV proteins by Log-rank test. Time represents the number of days from admission to time of onset for a severe COVID-19 related event. Optimal cut-off values were used to define high and low groups.

To further identify differences in EV proteins from Group 1 and Group 2 patients, a cross-validation score ^17^, which indicates the robustness of discrimination performance between them, was calculated on the basis of Fisher linear discriminant analysis for each of the selected proteins. From the candidate proteins, we have listed 91 proteins with cross-validation scores >0.75 (**Table S4**). Further, we show the abundance of the top 9 proteins with cross-validation scores >0.85 between the three patient groups (**Figure 1c**). Among these most-abundant proteins, four proteins [COPI coat complex subunit beta 2 (COPB2), KRAS proto-oncogene (KRAS), protein kinase C beta (PRKCB), and ras homolog family member C (RHOC)] are significantly more abundant in Group 1 than in Group 2 or in uninfected controls (*P* trend >0.05). Furthermore, CD147, calpain 2 (CAPN2), extracellular matrix protein 1 (ECM1), fibrinogen gamma chain (FGG) were significantly more abundant in Group 2 than in uninfected controls or in Group 1 (*P* trend <0.05). Only microfibril associated protein 4 (MFAP4) was less abundant in Group 1 than in uninfected controls or in Group 2 (*P* trend >0.05).

### Predictive value of 9 EV proteins for COVID-19 severity

We performed ROC analysis for combined Group1 and Group 2 to provide a robust test of sensitivity, specificity, and AUC for our set of nine predictive markers (**Figure 1d**). The AUC values for the four markers with enhanced abundance in Group 1, COPB2, KRAS, PRKCM, and RHOC, were 1.00 (95% CI: 1.00-1.00), 0.93 (95% CI: 0.85-1.00), 0.93 (95% CI: 0.83-1.00), and 0.96 (95% CI: 0.89-1.00), respectively. AUC values for the other five markers, CD147, CAPN2, ECM1, FGG, and MFAP4, were 0.73 (95% CI: 0.48-0.98), 0.84 (95% CI: 0.67-1.00), 0.82 (95% CI: 0.60-1.00), 0.87 (95% CI: 0.73-1.00), and 0.75 (95% CI: 0.52-0.97), respectively. Our analysis suggests that this set of markers, examined at the time of patient admission to the hospital, provides a significant degree of separation between patients that will develop mild versus severe COVID-19.

Additional ROC curves were generated to identify optimal cutoff values for the 9 marker proteins according to the Youden index (**Table S6**). For further analysis, we used the cutoff values to separate COVID-19 patients into low and high groups for comparing the onset of severe COVID-19 related events. Kaplan-Meier curves for the time-to-onset of a severe event were constructed for each of the 9 proteins (**Figure 1e**). Progression-free times for COPB2, KRAS, PRKCM, and RHOC, proteins with enhanced abundance in Group 1, were significantly shorter in the low group than in the high group (*P*=9.8×10^−10^; *P*=1.0×10^−5^; *P*=5.1×10^−7^; *P*=4.2×10^−8^, respectively). Conversely, the progression-free times for CD147, CAPN2, ECM1, FGG, and MFAP4, proteins with enhanced abundance in Group 2, were significant shorter in the high group than in the low group (*P*=7.0×10^−5^; *P*=0.00060; *P*=2.2×10^−5^; *P*=1.9×10^−7^; *P*=2.0×10^−6^, respectively). These results reinforce the concept that this set of markers can be valuable for predicting the likelihood of patients experiencing severe COVID-19 related events.

### NGS determination of ExRNA profiles in serum samples from COVID-19 patients and uninfected controls

Circulating exRNAs have the potential to serve as biomarkers for a wide range of diseases. ExRNAs consist of diverse RNA subpopulations that are protected from degradation by incorporation into EVs or by association with lipids and/or proteins. ExRNA profiles in blood samples are dynamic and include mRNAs, miRNAs, piRNAs, and lncRNAs ^18^. Here we have used next-generation sequencing (NGS) to analyze exRNAs present in patient serum samples (**Figure 2a**). From our 41 serum samples, NGS analysis identified 408 transcripts, excluding transcripts with fewer than 50 reads in all samples. Of these exRNAs, 43 transcripts were differentially expressed between the three patient groups (*P*<0.05; one-way ANOVA). To identify patterns of expression for these 43 transcripts among the three groups, we performed unsupervised multivariate statistics based on PCA mapping. PCA plots from the NGS data revealed a trend toward separation between the three groups (**Figure 2b**). This separation could be described by the first PC1, which accounted for 28.1% of the variance. The second PC2 accounted for 14.5% of the variance. These observations indicate that the serum exRNA profiles in COVID-19 patients deviate considerably from those in uninfected donors. Moreover, despite some overlap or dispersity, the PCA plots allow us to detect obvious differences in exRNA profiles between Group 1 and Group 2.

**Figure 2.**
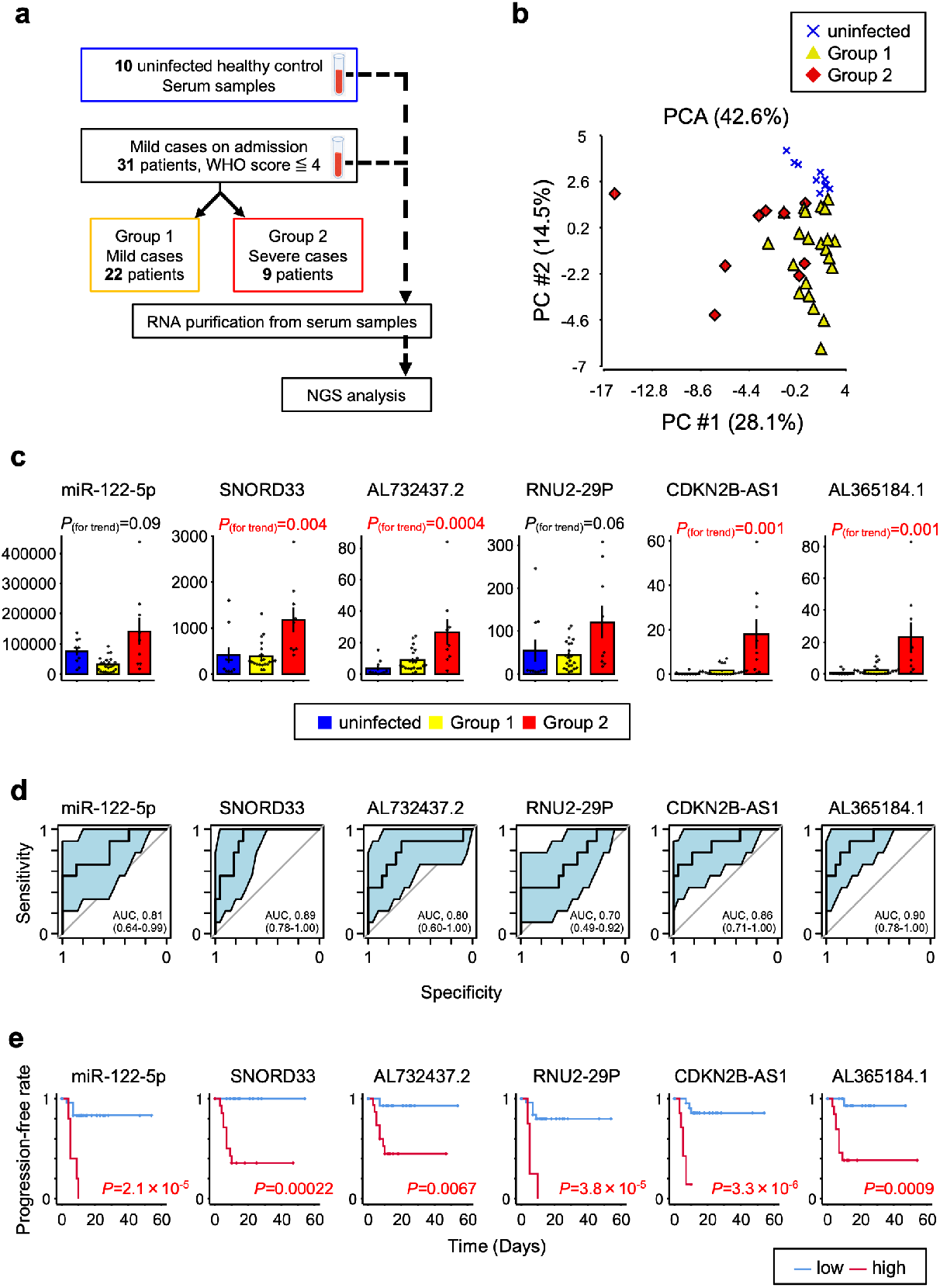
ExRNA profiles for early prediction of COVID-19 severity. (**a**) Work flow for NGS determination of exRNA profiles from serum samples of 31 mild COVID-19 patients (Group 1: *n*=22, Group2: *n*=9) and 10 uninfected healthy controls. (**b**) PCA map of 43 transcripts for the three subject groups. (**c**) Correlations of miR-122-5p, SNORD33, AL732437.2, RNU2-29P, CDKN2B-AS1, and AL365184.1 between the three subject groups. *P* values for trend by *Pearson’s* correlation analysis. Error bars represent mean + SEM. (**d**) AUC values (95% CI) for 6 transcripts evaluated by ROC analysis. (**e**) Kaplan-Meier curves for 6 transcripts by Log-rank test. Time represents the number of days from admission to time of onset for a severe COVID-19 related event. Optimal cut-off values were used to define high and low groups.

For discrimination between Group 1 and Group 2, a cross-validation score was calculated for each of the selected transcripts on the basis of Fisher linear discriminant analysis, much as described for our analysis of EV proteomes. From the candidate transcripts, we chose 14 transcripts with cross-validation scores >0.75 (**Table S5**). **Figure 2c** compares the three patient groups for expression of the top 6 transcripts with cross-validation scores>0.80. These species include miR-122-5p, small nucleolar RNA C/D Box 33 (SNORD33), AL732437.2, RNA U2 small nuclear 29 Pseudogene (RNU2-29P), CDKN2B antisense RNA1 (CDKN2B-AS1), and AL365184.1 (this transcript has 5 different transcript IDs). Notably, the four transcripts SNORD33, AL732437.2, CDKN2B-AS1, and AL365184.1 exhibit significantly higher levels of expression in Group 2 than in uninfected controls or Group 1 (*P* trend <0.05).

### Predictive value of 6 exRNAs for COVID-19 severity

Next, we constructed ROC curves for Group1 and Group 2 to provide a robust test of sensitivity, specificity, and AUC values for our set of 6 predictive exRNA markers (**Figure 2d**). AUC values for the transcripts miR-122-5p, SNORD33, AL732437.2, RNU2-29P, CDKN2B-AS1, and AL365184.1 were 0.81 (95% CI: 0.64-0.99), 0.89 (95% CI: 0.78-1.00), 0.80 (95% CI: 0.60-1.00), 0.70 (95% CI: 0.49-1.92), 0.86 (95% CI: 0.71-1.00), and 0.90 (95% CI: 0.78-1.00), respectively. This AUC analysis indicates that these exRNAs can provide a good basis for discriminating between mild and severe COVID-19 cases at the time of patient admission.

Additional ROC curves were generated to identify optimal cutoff values for the 6 transcripts according to the Youden index (**Table S6**). Using these cutoff values, we separated COVID-19 patients into low and high groups for determining the incidence of severe COVID-19 related events. Kaplan-Meier curves for time-to-onset of a severe event after admission were analyzed for each of the 6 transcripts (**Figure 2e**). The progression-free times for miR-122-5p, SNORD33, AL732437.2, RNU2-29P, CDKN2B-AS1, and AL365184.1 were all significantly faster in the high group than in the low group (*P*=2.1×10^−5^; *P*=0.00022; *P*=0.0067; *P*=3.8×10^−5^; *P*=3.3×10^−6^; *P*=0.0009, respectively). This finding suggests that these exRNA markers have value for predicting the incidence of severe COVID-19 related events at the time of patient admission to the hospital.

### The correlation between the selected markers for predicting disease severity values

Next we used univariate Cox regression analysis to calculate the hazard ratio (HR) for each of the EV and exRNA markers. Notably, the HR for COBP2 low was statistically incalculable using the optimal cut-off value, suggesting that EV COPB2 has the best predictive value among the two sets of markers. Furthermore, age high (HR 28.1; 95% CI 3.4-231.9; *P*=0.0019), CRP high (HR 8.4; 95% CI 1-67.5; *P*=0.045), PRKCB low (HR 32.1; 95% CI 3.9-261.9; *P*=0.0012), RHOC low (HR 23.6; 95% CI 4.7-118; *P*=0.00012), CD147 high (HR 10.7; 95% CI 2.5-45.1; *P*=0.0013), CAPN2 high (HR 15.5; 95% CI 1.9-125.9; *P*=0.010), ECM1 high (HR 11.6; 95% CI 2.8-48.4; *P*=0.00079), FGG high (HR 21.4; 95% CI 4.2-110.4; *P*=0.00025), MFAP4 high (HR 2.7; 95% CI 3.3-48.6; *P*=0.00022), miR-122-5p high (HR 10.5; 95% CI 2.7-40.4; *P*=0.00063), AL732437.2 high (HR 9.9; 95% CI 1.2-79.9; *P*=0.031), RNU2-29P high (HR 10.4; 95% CI 2.6-40.8; *P*=0.00081), CDKN2B-AS1 high (HR 14.4; 95% CI 3.4-61.3; *P*=0.00031), and AL365184.1 high (HR 14.2; 95% CI 1.8-114.4; *P*=0.013) were statistically significant (**Table S6**).

To investigate potential relationships between the selected markers, *Spearman’s* correlation coefficients were calculated based on marker levels. A correlation plot was constructed to visualize the correlation coefficients of the 19 markers (**Figure 3)**. This allowed identification of four hierarchical clusters of markers that share strong positive correlations within the groups to which they belong. Each marker appears to fit into one of the four well defined clusters (namely, cluster 1, 2, 3, and 4). Notably, we observe that cluster 1 (PRKCB, RHOC, COPB2, and KRAS) is negatively correlated with the other clusters. Clusters 2, 3, and 4 have substantially strong positive correlations with each other.

**Figure 3.**
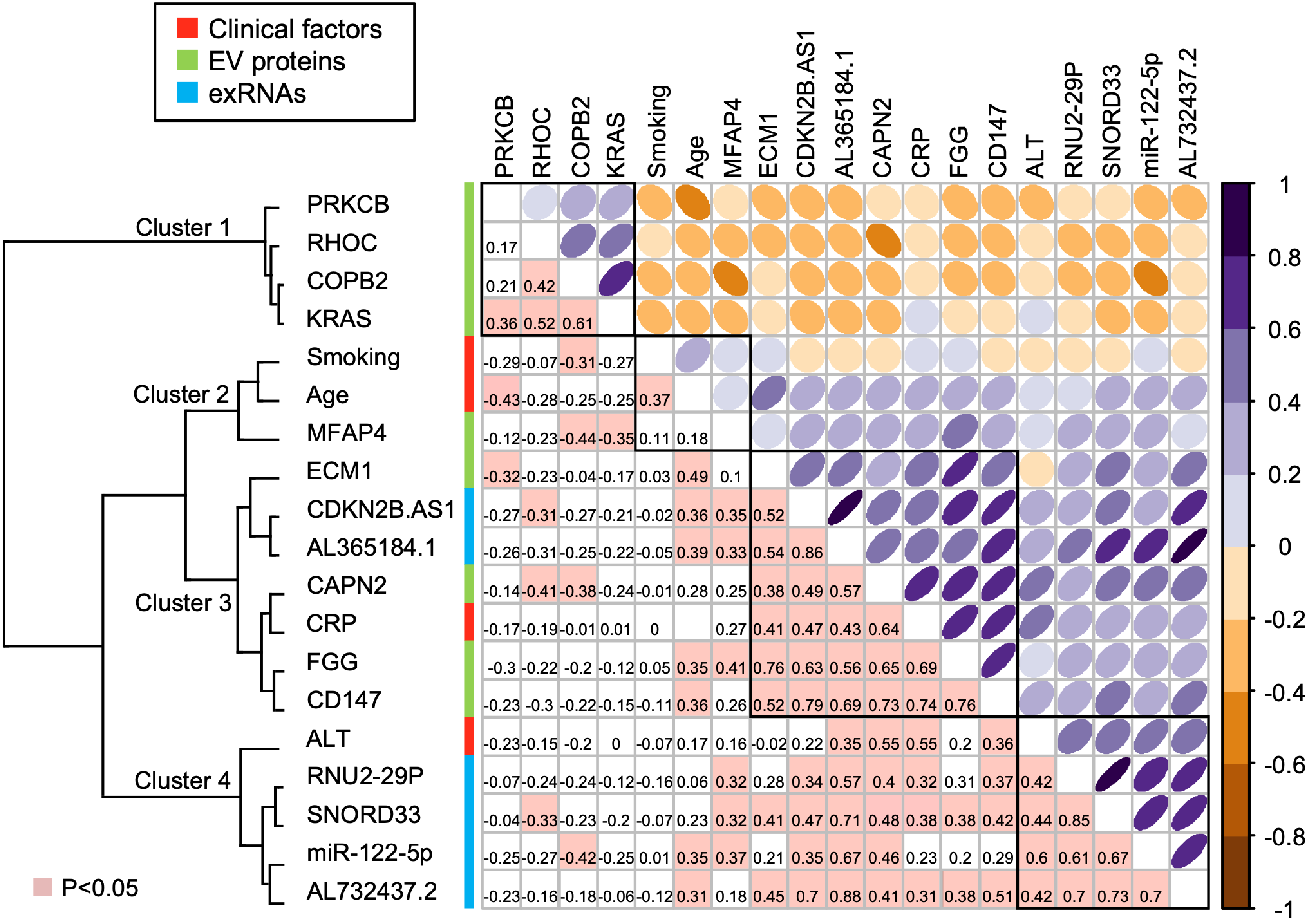
Upper triangular correlation plot of the associations between 4 clinical factors, 9 EV proteins, and 6 transcripts. Colors represent Pearson’s correlation coefficients. Positive correlations are represented in purple, while negative correlations are represented in brown in the upper triangle. Color intensity and ovalization of the circle are proportional to the correlation coefficients. The lower triangular correlation matrix displays actual correlation values, with pink highlights representing *P*<0.05. Cluster 1 (PRKCB, RHOC, COPB2, and KRAS) contains a group of antiviral response-related EV proteins. Clusters 2 (smoking, age, and MFPA4) and 3 (CM1, CDKN2B.AS1, AL365184.1, CAPN2, CRP, FGG, and CD147) contain groups of coagulation-related markers. Cluster 4 (ALT, RNU2-29P, SNORD33, miR-122-5p, and AL732437.2) contains a group of liver damage-related exRNAs.

All 4 EV proteins in cluster 1 exhibit significantly higher abundance in Group 1 than in Group 2 COVID-19 patients. This is consistent with the idea that cluster 1 might represent a group of antiviral response-related EV proteins. Although the functions of RHOC or KRAS during SARS-CoV-2 infection remain unknown, there are some reports that implicate PRKCB or COPB2 in antiviral effects against SARS-CoV-2. PRKCB can regulate metabolic and mitochondrial aspects of reprogramming responsible for B cell fate ^19^. A recent bioinformatics-based report has revealed that PRKCB is one of the target genes activated by vitamin A that may have antiviral potential against SARS-CoV-2 ^20^. COPB2 is a subunit of the Golgi Coatomer complex (COPI) that is necessary for retrograde trafficking from the Golgi to the endoplasmic reticulum (ER). Many viruses, including RNA viruses, DNA viruses, and retroviruses, hijack or adapt COPI-related proteins for their own benefit. Indeed, the gene is required for replication of SARS-CoV-1, which is closely genetically related (79% identity) to SARS-CoV-2 ^21^. Depletion of COPB2 has a strong antiviral effect, based on reduced SARS-CoV-1 protein expression ^22^. Although the precise functions of COPB2 positive EVs still remain unknown, we hypothesize that secretion of these EVs reflects the development of patient antiviral responses against SARS-CoV-2, potentially mitigating the severity of COVID-19 progression.

Cluster 2 includes smoking, age, and MFPA4. MFAP4 is an extracellular matrix protein belonging to the fibrinogen-related protein superfamily ^23^. Levels of MFAP4 are not significantly correlated with either smoking or age. On the other hand, cluster 3 includes ECM1, CDKN2B.AS1, AL365184.1, CAPN2, CRP, FGG, and CD147. Based on previous evidence, we considered that ECM1, CDKN2B.AS1, CAPN2, and CD147 could be implicated in the abnormal coagulation status associated with severe COVID-19. The extracellular matrix protein ECM1 is expressed in association with blood vessels and may have pro-angiogenic functions ^24^. The long non-coding RNA CDKN2B.AS1 regulates production of extracellular matrix in the pathogenesis associated with complications of diabetes 25. CAPN2 is a calcium-dependent intracellular protease that maintains the level of matrix metalloproteinase 2, helping to regulate extracellular matrix accumulation ^26^. CD147, known as extracellular matrix metalloproteinase inducer (EMMPRIN), is a transmembrane glycoprotein considered to be a binding partner for the SARS-CoV-2 spike protein, with obvious functional implications for viral infection ^27^. Furthermore, the role of CD147 in matrix metalloproteinase regulation is important for tumor progression and for development of coronary artery disease, affecting both plaque stability and thrombus formation ^28^. The levels of 1 exRNA (CDKN2B.AS1 from cluster 3) and 4 proteins relevant to extracellular matrix formation (MFPA4 from cluster 2 and ECM1, CAPN2, and CD147 from cluster 3) are correlated with the levels of FGG, which has crucial functions in coagulation (*P*<0.05) (**Figure 3**). Reports of widespread thromboses and disseminated intravascular coagulation (DIC) in COVID-19 patients have been rapidly increasing in number. Recent reports show that D-dimer elevation at the time of admission is predictive of bleeding, thrombosis, critical illness, and death in COVID-19 patients ^29^. Unfortunately, as our study includes only one COVID-19 patient with complications from thrombosis, we are unable to make correlations between D-dimer elevation and predicted disease severity. With regard to the relationship between cluster 2 and 3, the levels of markers in cluster 3 are correlated with age, which in turn is related to vascular endothelial dysfunction and coagulation ^30^. Although the function of non-coding RNA AL365184.1 is unknown, the bulk of our data suggest that cluster 2 and 3 represent groups of coagulation-related markers.

The Cluster 4 components ALT, RNU2-29P, SNORD33, miR-122-5p, and AL732437.2 may at least partly reflect phenomena related to liver damage. Levels of ALT, a representative transaminase mainly associated with liver dysfunction, correlate with levels of the three exRNA species (*P*<0.05). miRNA-122-5p in particular is a promising exploratory biomarker for detecting liver injury in preclinical and clinical studies ^31^. It has been reported that the small nucleolar RNA SNORD33 is induced in response to liver injury caused by lipopolysaccharide (LPS) ^32^. LPS-induced inflammation has also been found to stimulate secretion of SNORD33 packaged into EVs ^33^. Recent data indicate that deranged expression of liver enzymes is one manifestation of COVID-19 pathology, and that liver injury is more prevalent in severe cases than in mild cases of COVID-19 ^34,35^. Although the functions of the non-coding RNAs RNU2-29P and AL732437.2 are unknown, our data at least partially support the idea that exRNAs associated with liver damage can serve as biomarkers for predicting the severity of COVID-19 in patients at the time of admission.

## Discussion

As a single center study with a small sample size, our work has several limitations, and caution should be exercised in utilizing the predictive value of the markers we have identified. An expanded random sample across all genders and ages should be more representative of the general population, and larger clinical studies are required to validate the potential of these biomarkers for predicting the severity of COVID-19 progression. Nevertheless, our research has implications that go beyond the simple investigation of biomarkers, since the results also provide important clues regarding the pathogenesis of COVID-19 and the development of therapies for the disease. Indeed, biomarkers such as PRKCB, COPB2, and CD147 may be involved in SARS-CoV-2 infection or replication. Furthermore, 6 markers (MFAP4, ECM1, CDKN2B.AS1, CAPN2, FGG, and CD147) and 2 exRNAs (SNORD33 and miR-122-5p) are involved in thrombosis or liver injury, which are serious complications in patients with severe COVID-19. These findings indicate that the profiles of EV proteins and exRNAs in patient sera clearly reflect specific host reactions to SARS-CoV-2 infection and progression of the disease. Although the pathological significance of some markers is unknown, understanding the profiles of functional extracellular components in patient sera may help clarify various aspects of COVID-19 pathogenesis. With regard to therapy, biomarkers may provide information concerning potential therapeutic targets for mitigating the effects of SARS-CoV-2 infection. For instance, our finding that 4 EV proteins in cluster 1 are significantly more abundant in Group 1 than in Group 2 suggests that these proteins could have antiviral effects. Supplementation with these EVs could protect patients against COVID-19 progression and the associated complications. In addition, a recent study showed that antibody against the spike protein receptor CD147 could block infection by SARS-CoV-2 ^27^. Similarly, neutralization or manipulation of other markers with higher abundance in Group 2 than in Group 1 might also have antiviral effects or provide protection against severe consequences of COVID-19 progression.

In summary, our comprehensive evaluation identifies 3 distinct groups of components (antiviral response-related EV proteins, coagulation-related markers, and liver damage-related exRNAs) capable of serving as early predictive biomarkers for the severity of COVID-19 progression. Among these markers, EV COPB2 has the best predictive value for severe deterioration of COVID-19 patients in our cohort. COVID-19 patients with high levels of EV COPB2 at the time of admission might be able to overcome the disease without experiencing severe events. It might be worthwhile to consider whether these markers have greater predictive power than simpler factors like age and CRP levels. This study for the first time provides potential resources for early discrimination between COVID-19 patients that may be resistant to disease progression and patients that are likely to experience severe COVID-19 related deterioration. Our results also suggest that, in addition to their predictive value, functional extracellular components can also be potential contributors and mitigators of pathogenesis during COVID-19 deterioration. These ideas should be validated and expanded in future studies.

## Methods

### Study approval

This retrospective study involving collection of COVID-19 serum samples was approved by the Institutional Review Board at The Jikei University School of Medicine (Number: 32-055(10130)). The protocol did not require informed consent, and patients were given the choice of opting out of the study. For collection of healthy control serum samples, this study was approved by the Institutional Review Board at The Institute of Medical Science, The University of Tokyo (Number: 28-19-0907). Written informed consent was provided by all healthy donors before sample acquisition, in accordance with Declaration of Helsinki principles.

## Data Availability

GSE data will be opened.

## Aknowledgement

We thank Yusuke Hosaka and Akihiko Ito (The Jikei University School of Medicine, Tokyo, Japan), Dr. Yoshihiro Hirata (The Institute of Medical Science, The University of Tokyo), and Dr. Takashi Nakagawa (Omiya City Clinic) for clinical sample collection, Dr Misato Yamamoto for technical assistance, Dr. Tatsutoshi Inuzuka for technical support of LC-MS analysis (H.U. Group Research Institute), and all medical staff of Team COVID at The Jikei University Hospital. This work was supported by International Space Medical Co., Ltd.

## Author contributions

Y.F. and T.O. conceived the idea and coordinated the project. Y.F., T.Hoshina., J.M., T.K., S.F., H.K., and N.W. performed statistical data analysis, and wrote the draft of the manuscript. T.Hoshina., K.S., Y.S., M.Miyajima., K.L., K.N., T.Horino., R.N., and M.Y. collected serum samples. M.Miyato., J.A., and K.K. provided helpful discussion. The manuscript was finalized by Y.F. with the assistance of all authors.

## References

1. Dong, E., Du, H. & Gardner, L. An interactive web-based dashboard to track COVID-19 in real time. Lancet Infect Dis 20, 533–534 (2020).

2. Guan, W.J., et al. Clinical Characteristics of Coronavirus Disease 2019 in China. N Engl J Med 382, 1708–1720 (2020).

3. Huang, C., et al. Clinical features of patients infected with 2019 novel coronavirus in Wuhan, China. Lancet 395, 497–506 (2020).

4. Huang, D., et al. Clinical features of severe patients infected with 2019 novel coronavirus: a systematic review and meta-analysis. Ann Transl Med 8, 576 (2020).

5. Mangalmurti, N. & Hunter, C.A. Cytokine Storms: Understanding COVID-19. Immunity 53, 19–25 (2020).

6. Chen, N., et al. Epidemiological and clinical characteristics of 99 cases of 2019 novel coronavirus pneumonia in Wuhan, China: a descriptive study. Lancet 395, 507–513 (2020).

7. Yanez-Mo, M., et al. Biological properties of extracellular vesicles and their physiological functions. J Extracell Vesicles 4, 27066 (2015).

8. Kowal, J., et al. Proteomic comparison defines novel markers to characterize heterogeneous populations of extracellular vesicle subtypes. Proc Natl Acad Sci U S A 113, E968–977 (2016).

9. Robbins, P.D. & Morelli, A.E. Regulation of immune responses by extracellular vesicles. Nat Rev Immunol 14, 195–208 (2014).

10. Pegtel, D.M., et al. Functional delivery of viral miRNAs via exosomes. Proc Natl Acad Sci U S A 107, 6328–6333 (2010).

11. Earnest, J.T., et al. The tetraspanin CD9 facilitates MERS-coronavirus entry by scaffolding host cell receptors and proteases. PLoS Pathog 13, e1006546 (2017).

12. Wang, J., Chen, S. & Bihl, J. Exosome-Mediated Transfer of ACE2 (Angiotensin-Converting Enzyme 2) from Endothelial Progenitor Cells Promotes Survival and Function of Endothelial Cell. Oxid Med Cell Longev 2020, 4213541 (2020).

13. Das, S., et al. The Extracellular RNA Communication Consortium: Establishing Foundational Knowledge and Technologies for Extracellular RNA Research. Cell 177, 231–242 (2019).

14. enOever, B.R. RNA viruses and the host microRNA machinery. Nat Rev Microbiol 11, 169–180 (2013).

15. Girardi, E., Lopez, P. & Pfeffer, S. On the Importance of Host MicroRNAs During Viral Infection. Front Genet 9, 439 (2018).

16. Wang, Z., Yang, B., Li, Q., Wen, L. & Zhang, R. Clinical Features of 69 Cases With Coronavirus Disease 2019 in Wuhan, China. Clin Infect Dis 71, 769–777 (2020).

17. Urabe, F., et al. Large-scale Circulating microRNA Profiling for the Liquid Biopsy of Prostate Cancer. Clin Cancer Res 25, 3016–3025 (2019).

18. Murillo, O.D., et al. exRNA Atlas Analysis Reveals Distinct Extracellular RNA Cargo Types and Their Carriers Present across Human Biofluids. Cell 177, 463–477 e415 (2019).

19. Tsui, C., et al. Protein Kinase C-beta Dictates B Cell Fate by Regulating Mitochondrial Remodeling, Metabolic Reprogramming, and Heme Biosynthesis. Immunity 48, 1144–1159 e1145 (2018).

20. Li, R., et al. Revealing the targets and mechanisms of vitamin A in the treatment of COVID-19. Aging (Albany NY) 12, 15784–15796 (2020).

21. Lu, R., et al. Genomic characterisation and epidemiology of 2019 novel coronavirus: implications for virus origins and receptor binding. Lancet 395, 565–574 (2020).

22. de Wilde, A.H., et al. A Kinome-Wide Small Interfering RNA Screen Identifies Proviral and Antiviral Host Factors in Severe Acute Respiratory Syndrome Coronavirus Replication, Including Double-Stranded RNA-Activated Protein Kinase and Early Secretory Pathway Proteins. J Virol 89, 8318–8333 (2015).

23. Wulf-Johansson, H., et al. Localization of microfibrillar-associated protein 4 (MFAP4) in human tissues: clinical evaluation of serum MFAP4 and its association with various cardiovascular conditions. PLoS One 8, e82243 (2013).

24. Steinhaeuser, S.S., et al. ECM1 secreted by HER2-overexpressing breast cancer cells promotes formation of a vascular niche accelerating cancer cell migration and invasion. Lab Invest 100, 928–944 (2020).

25. Thomas, A.A., Feng, B. & Chakrabarti, S. ANRIL regulates production of extracellular matrix proteins and vasoactive factors in diabetic complications. Am J Physiol Endocrinol Metab 314, E191–E200 (2018).

26. Jang, H.S., Lal, S. & Greenwood, J.A. Calpain 2 is required for glioblastoma cell invasion: regulation of matrix metalloproteinase 2. Neurochem Res 35, 1796–1804 (2010).

27. Aguiar, J.A., et al. Gene expression and in situ protein profiling of candidate SARS-CoV-2 receptors in human airway epithelial cells and lung tissue. Eur Respir J 56(2020).

28. Joghetaei, N., et al. The Extracellular Matrix Metalloproteinase Inducer (EMMPRIN, CD147) - a potential novel target in atherothrombosis prevention? Thromb Res 131, 474–480 (2013).

29. Al-Samkari, H., et al. COVID-19 and coagulation: bleeding and thrombotic manifestations of SARS-CoV-2 infection. Blood 136, 489–500 (2020).

30. Donato, A.J., Machin, D.R. & Lesniewski, L.A. Mechanisms of Dysfunction in the Aging Vasculature and Role in Age-Related Disease. Circ Res 123, 825–848 (2018).

31. Bala, S., et al. Circulating microRNAs in exosomes indicate hepatocyte injury and inflammation in alcoholic, drug-induced, and inflammatory liver diseases. Hepatology 56, 1946–1957 (2012).

32. Michel, C.I., et al. Small nucleolar RNAs U32a, U33, and U35a are critical mediators of metabolic stress. Cell Metab 14, 33–44 (2011).

33. Rimer, J.M., et al. Long-range function of secreted small nucleolar RNAs that direct 2’-O-methylation. J Biol Chem 293, 13284–13296 (2018).

34. Zhang, C., Shi, L. & Wang, F.S. Liver injury in COVID-19: management and challenges. Lancet Gastroenterol Hepatol 5, 428–430 (2020).

35. Hajifathalian, K., et al. Gastrointestinal and Hepatic Manifestations of 2019 Novel Coronavirus Disease in a Large Cohort of Infected Patients From New York: Clinical Implications. Gastroenterology 159, 1137–1140 e1132 (2020).

