## Supplementary data for "Early Prediction of COVID-19 Severity Using Extracellular Vesicles and Extracellular RNAs"

**Supplementary Informations**

### Methods

#### Study subjects

We enrolled 42 patients positive for SARS-CoV-2 who were admitted to the Jikei University Hospital between March and May 2020 (approved by the Institutional Review Board at The Jikei University School of Medicine (Number: 32-055(10130)). COVID-19 patients were recruited on the basis of positive PCR tests for SARS-CoV-2 RNA obtained by nasopharyngeal swabs. The eventual degree of COVID-19 severity was categorized as mild or severe on the basis of WHO 2020 scoring (**Table S2**). 11 Patients with severe COVID-19 at admission were excluded. At the time of admission, all COVID-19 patients were scored as having a mild status (WHO score = 3). Based on the clinical disease course after admission, we divided 31 patients in two groups (namely, Group 1: mild cases (WHO score ≦ 4), and Group 2: severe cases (WHO score ≧ 5)). All patients received standard therapy without corticosteroids according to Clinical management of COVID-19 interim guidance by WHO. 10 healthy age-matched donors were recruited at Omiya City Clinic, Saitama for routine medical examination between March and April 2019 (approved by the Institutional Review Board at The Institute of Medical Science, The University of Tokyo (Number: 28-19-0907). The medical records of COVID-19 patients and healthy donors were analyzed retrospectively. 31 serum samples taken at admission from COVID-19 patients and 10 serum samples taken from healthy donors were separated by centrifugation at 3000 rpm for 10 min at 4°C. The supernatant was collected into a new tube and the serum sample was stored at -80°C until use.

#### Isolation of EVs

Anti-CD9 antibody and anti-CD63 antibody (H.U. Group Research Institute, Tokyo) coupled to Dynabeads M-280 Tosylactivated (Thermo Fisher Scientific Inc, Waltham, MA, USA) were added to aliquots (500 μL) of the serum samples that had been treated with chelate-based PEVIA® reagent (H.U. Group Research Institute), followed by incubation on a rotator at 4°C for 18 h. The beads were washed three times with PBS and stored at 4°C until further analysis.

#### Preparation of peptides

EVs were processed using S-Trap micro spin columns (AMR Inc, Tokyo, Japan) according to the manufacturer's instruction with minor modifications. In brief, captured EVs were suspended in 50 µL of 5% SDS (FUJIFILM Wako Pure Chemical Corporation, Osaka, Japan) in 50mM TEAB buffer (Honeywell Inc, Charlotte, NC, USA), pH7.5. After removing beads, the amount of protein from EVs were determined by Micro BCA™ Protein Assay Kit (Thermo Fisher Scientific Inc). 13.8 ng Pierce™ Digestion Indicator for Mass Spectrometry (Thermo Fisher Scientific Inc) was added to lysed samples for quality control of digestion efficiency. The samples were then reduced and alkylated with dithiothreitol (FUJIFILM Wako Pure Chemical Corporation) and iodoacetamide (Nacalai tesque Inc, Kyoto, Japan), respectively. 12% aqueous phosphoric acid (FUJIFILM Wako Pure Chemical Corporation) was added to a final concentration of 1.2% followed by six times the volume of S-Trap protein binding buffer. The sample mixtures were added to S-Trap columns that were prewashed and preconditioned with 0.2 % formic acid in 50 % acetonitrile and S-Trap buffer, respectively. The S-Trap columns were washed with 150uL S-Trap buffer. Centrifugation and removal of the flow through were then repeated 6 times. Washed columns were incubated with 20uL of digestion buffer containing 0.75ug Trypsin/Lys-C Mix, Mass Spec Grade (Promega Corporation, Madison, WI, USA) for 2hr at 47°C. After digestion, peptides were eluted from the S-Trap column, lyophilized with miVac system (Genevac Ltd, Ipswich, United Kingdom), and stored at -80 °C until use.

**Proteomic analysis with LC-MS**

Peptides obtained from EV proteins were reconstituted in 10 µl of water containing 0.1% formic acid (FA) (Fisher Chemical, Thermo Fisher Scientific Inc). Quantification of peptides was accomplished using Pierce™ Quantitative Fluorometric Peptide Assay (Thermo Fisher Scientific Inc). Proteomic analysis of the peptides was carried out using Q Exactive (Thermo Fisher Scientific Inc.) equipped with UltiMate 3000 Nano LC Systems (Thermo Fisher Scientific Inc.). Peptide samples (1 µg) were injected onto Acclaim PepMap 1000 trap columns (75 µm × 2 cm, nanoViper C18 3 µm, 100Å, Thermo Fisher Scientific Inc) which were heated to 40 °C in a chamber which was connected to a C18 reverse-phase Aurora UHPLC Emitter Column with nano Zero & Captive Spray Insert (75 µm × 25 cm, Ion Opticks Pty Ltd) using Dreamspray interface (AMR Inc). The nano pump flow rate was set to 250 nL/min with a 302 min gradient, in which the mobile phases were A (0.1% FA in water, Fisher Chemical, Thermo Fisher Scientific Inc.) and

B (0.1% FA in acetonitrile, Fisher Chemical, Thermo Fisher Scientific Inc.). The chromatography gradient was designed to provide a linear increase from 0-8 min at 2% B, 8-272 min from 2% B to 35% B, 272-282 min from 35% B to 70% B, 282-283 min from 70% B to 95% B, wash, 8 min and 10 min equilibrium. The data-dependent acquisition was performed in positive ion mode. Mass spectrometry parameters and those of the Proteome Discoverer 2.2.0.388 software (Thermo Fisher Scientific Inc) were described in a previous report (doi: https://doi.org/10.1101/2020.06.17.155861).

**Analysis of serum exRNA profiles**

Total RNA was extracted from aliquots (200 μL) of the serum samples using QIAzol and the miRNeasy Mini Kit (Qiagen, Hilden, Germany) according to the manufacturer's protocol. The library was prepared using the QIAseq miRNA Library Kit (Qiagen). Library preparations were subjected to quality control using either a Bioanalyzer 2100 or TapeStation 4200 system (Agilent Technologies, Santa Clara, CA, USA). The library pools were quantified using the Library Quantification Kit (Takara, Shiga, Japan) and sequenced on the NovaSeq 6000 sequencing platform (Illumina Inc, San Diego, CA, USA). Reads were pre-processed and annotated against miRBase v22.1 and Ensembl non-coding RNA database release 100 by using CLC Genomics Workbench v20.0.1. Raw and normalized microarray data is available in the Gene Expression Omnibus database (GSE158877).

**Statistical analysis**

Fisher's exact test for categorical variables and unpaired Student's *t*-test for continuous variables were used to compare clinical data between two groups. To identify biomarker candidates among EV proteins and exRNAs, we initially used one-way analysis of variance (ANOVA) to select candidates present at different levels in the three subject groups (uninfected, COVID-19 Group 1, and Group 2) with $P < 0.05$. Principal component analysis (PCA) was performed with the selected candidates using Partek Genomics Suite 7.0 (Partek, St. Louis, MO, USA). Second, candidates with better discrimination between Group 1 and 2 were selected based on linear discriminant analysis with leave-one-out cross-validation, and subsequent ROC analysis was performed using R version 3.6.3 (R Foundation for Statistical Computing, http:// www.R-project.org), compute.es package version 0.2-2, hash package version 2.2.6.1, MASS package version 7.3-51.5, mutoss package version 0.1-12, and pROC package version 1.16.2. Optimal cut-off values for each candidate were set based on the maximum point of the sum of sensitivity and specificity (Youden index). Predictive sensitivity, specificity, and accuracy were calculated with the corresponding cut-off value for each candidate. Kaplan–Meier analysis with log-rank test and Cox regression analysis were performed using IBM SPSS Statistics 25 (IBM Japan, Tokyo, Japan). The correlation plot was generated using R version 3.6.3 and corrplot package version 0.84., and the unsupervised hierarchical clustering analysis was performed using Partek Genomics Suite 7.0. The limit of statistical significance for all analyses was defined as a two-sided P value of 0.05.

**Supplementary Data**

**Figure S1.** Clinical factors at admission for early prediction of COVID-19 severity.

**Table S1**. Baseline characteristics of healthy donors and COVID-19 patients.

**Table S2**. WHO 2020 scoring for COVID-19 cases used in this study.

**Table S3**. Differences in clinical characteristics among healthy donors and COVID-19 patients.

**Table S4**. EV proteins for discrimination between mild and severe COVID-19 patients.

**Table S5**. ExRNAs for discrimination between mild and severe COVID-19 patients.

**Table S6**. Univariate Cox regression analysis of the selected factors and biomarkers for predicting COVID-19 severity in this patient cohort.

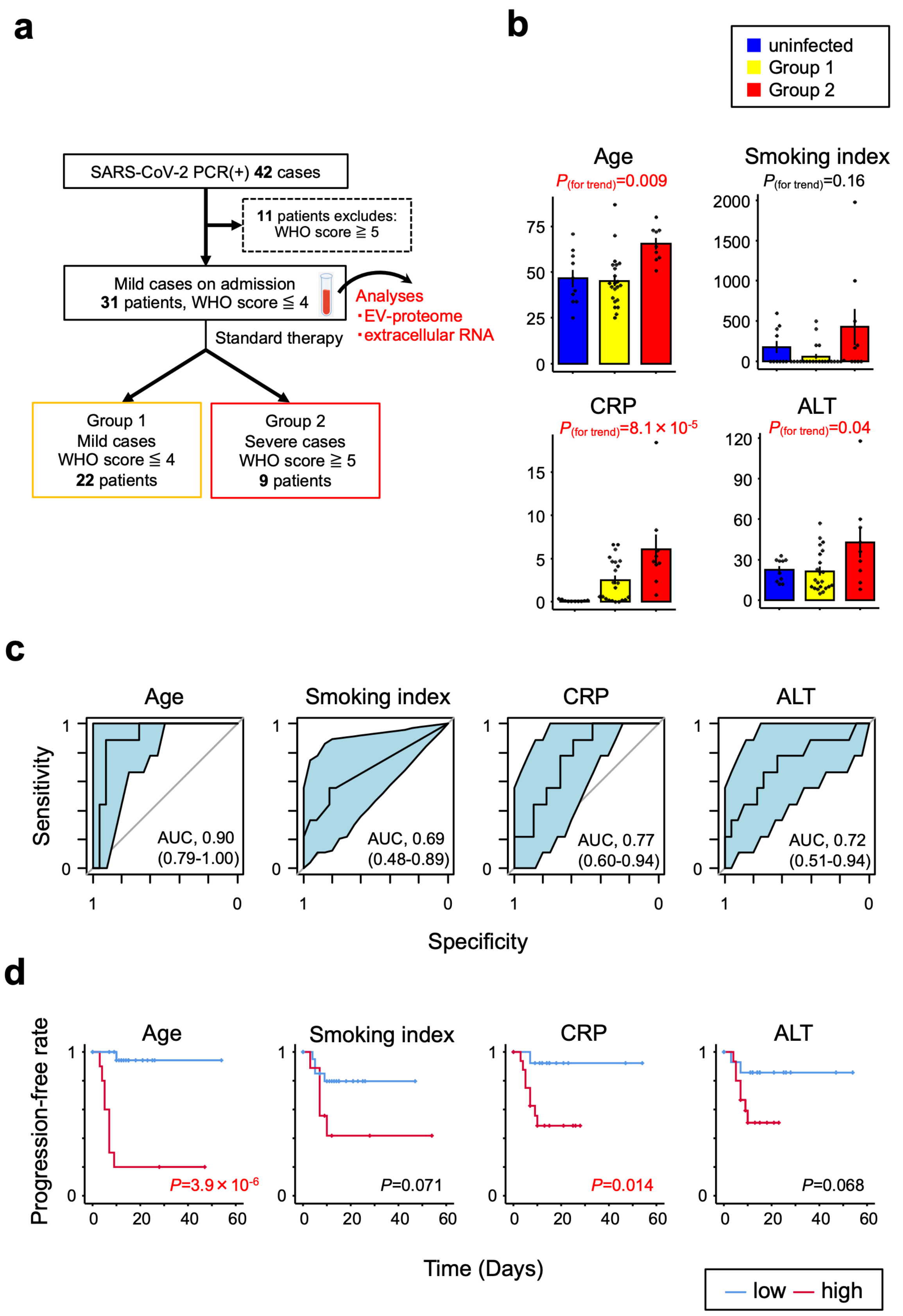


**Figure S1. Clinical factors at admission for early prediction of COVID-19 severity.** (**a**) Patient recruitment flowchart for this cohort. (**b**) Correlations of age, smoking index, CRP, and ALT between the three subject groups. *P* values for trend by *Pearson's* correlation analysis. Error bars represent mean ± SEM. (**c**) The AUC (95% CI) for age, smoking index, CRP, and ALT evaluated by ROC analysis. (**d**) Kaplan-Meier curves for age, smoking index, CRP, and ALT by Log-rank test. Time represents the number of days from admission to time of onset for a severe COVID-19 related event. In each case, optimal cut-off values were used to define high and low groups.

### Table S1. Baseline characteristics of healthy donors and COVID-19 patients.

| | Patients | WHO score | WHO category | Age (10-year range) | Sex | Onset of severe events (day)† | The period of hospital stay | Outcome |
|---|---|---|---|---|---|---|---|---|
| Healthy donor | HD-1 | 0 | uninfected | 30's | M | - | - | - |
| | HD-2 | 0 | uninfected | 30's | M | - | - | - |
| | HD-3 | 0 | uninfected | 20's | F | - | - | - |
| | HD-4 | 0 | uninfected | 40's | M | - | - | - |
| | HD-5 | 0 | uninfected | 70's | M | - | - | - |
| | HD-6 | 0 | uninfected | 50's | M | - | - | - |
| | HD-7 | 0 | uninfected | 60's | M | - | - | - |
| | HD-8 | 0 | uninfected | 40's | M | - | - | - |
| | HD-9 | 0 | uninfected | 50's | M | - | - | - |
| | HD-10 | 0 | uninfected | 30's | F | - | - | - |
| Group 1 COVID-19 | 1 | 3→3 | mild→mild | 30's | M | - | 15 | discharged |
| | 2 | 3→3 | mild→mild | 30's | M | - | 14 | discharged |
| | 3 | 3→3 | mild→mild | 40's | M | - | 10 | discharged |
| | 4 | 3→3 | mild→mild | 40's | M | - | 13 | discharged |
| | 5 | 3→3 | mild→mild | 50's | M | - | 21 | discharged |
| | 6 | 3→3 | mild→mild | 50's | M | - | 26 | discharged |
| | 7 | 3→3 | mild→mild | 80's | M | - | 28 | discharged |
| | 8 | 3→3 | mild→mild | 50's | F | - | 25 | discharged |
| | 9 | 3→3 | mild→mild | 20's | F | - | 11 | discharged |
| | 10 | 3→3 | mild→mild | 20's | F | - | 23 | discharged |
| | 11 | 3→3 | mild→mild | 40's | F | - | 15 | discharged |
| | 12 | 3→3 | mild→mild | 40's | M | - | 12 | discharged |
| | 13 | 3→3 | mild→mild | 70's | M | - | 47 | discharged |
| | 14 | 3→3 | mild→mild | 40's | M | - | 54 | discharged |
| | 15 | 3→3 | mild→mild | 40's | M | - | 9 | discharged |
| | 16 | 3→3 | mild→mild | 50's | M | - | 15 | discharged |
| | 17 | 3→3 | mild→mild | 30's | F | - | 12 | discharged |
| | 18 | 3→3 | mild→mild | 50's | M | - | 18 | discharged |
| | 19 | 3→3 | mild→mild | 40's | F | - | 21 | discharged |
| | 20 | 3→3 | mild→mild | 30's | M | - | 0 | to medical facility |
| | 21 | 3→3 | mild→mild | 30's | F | - | 0 | to medical facility |
| | 22 | 3→3 | mild→mild | 40's | M | - | 7 | discharged |
| Group 2 COVID-19 | 23 | 3→7 | mild→ severe | 60's | M | 5 | 93 | discharged |
| | 24 | 3→7 | mild→ severe | 80's | M | 4 | 23 | death (pulmonary embolism) |
| | 25 | 3→7 | mild→ severe | 60's | M | 3 | 138 | still hospitalized |
| | 26 | 3→7 | mild→ severe | 70's | M | 9 | 44 | discharged |
| | 27 | 3→7 | mild→ severe | 70's | M | 7 | 29 | discharged |
| | 28 | 3→7 | mild→ severe | 70's | M | 7 | 125 | still hospitalized |
| | 29 | 3→7 | mild→ severe | 50's | M | 10 | 43 | death (ARDS) |
| | 30 | 3→6 | mild→ severe | 50's | M | 5 | 31 | discharged |
| | 31 | 3→5 | mild→ severe | 50's | M | 7 | 16 | discharged |

M, male; F, female; † Day after sampling point on admission.

### Table S2. WHO 2020 scoring for COVID-19 cases used in this study.

| Patient | Descriptor | Score |
|---|---|---|
| Uninfected | No clinical or virological evidence of infection | 0 |
| Ambulatory | No limitation of activities | 1 |
| | Limitation of activities | 2 |
| Hospitalized-mild disease | No oxygen therapy | 3 |
| | Oxygen by nasal prongs or mask | 4 |
| Hospitalized-severe disease | Non-invasive ventilation of high-flow oxygen | 5 |
| | Intubation and mechanical ventilation | 6 |
| | Ventilation + additional organ support (pressors, RRT, ECMO) | 7 |

RRT, renal replacement therapy; ECMO, extracorporeal membrane oxygenation.

### Table S3. Differences in clinical characteristics among healthy donors and COVID-19 patients.

| | **uninfected** (n=10) | **Group 1 COVID-19** (n=22) | **Group 2 COVID-19** (n=9) | ***P* value** | |
|---|---|---|---|---|---|
| | | | | (uninfected vs. infected) | (Group 1 vs. Group 2) |
| Age (year) | 46.5 ± 14.8 | 45.3 ± 14.3 | 65.4 ± 9.5 | 0.42 | 0.001 |
| Sex | | | | | |
| Men | 8 (80%) | 15 (68.2%) | 9 (100%) | 1.00 | 0.08 |
| Women | 2 (20%) | 7 (31.8%) | 0 (0%) | | |
| BMI ($kg/m^2$) | 22.8 ± 4.4 | 23.0 ± 3.0 (n=20) | 22.4 ± 3.2 | 0.95 | 0.63 |
| Smoking index | 176 ± 237 | 60 ± 140 | 427 ± 670 | 0.94 | 0.02 |
| WBC count ($10^3$/μL) | 6.7 ± 2.9 | 4.6 ± 1.5 | 5.2 ± 3.2 | 0.03 | 0.46 |
| CRP (mg/dL) | 0.12 ± 0.12 | 2.50 ± 2.41 | 6.07 ± 5.11 | 0.01 | 0.01 |
| BUN (mg/dL) | 13.7 ± 4.5 | 17.0 ± 16.6 | 22.6 ± 14.6 | 0.35 | 0.39 |
| Cr (mg/dL) | 0.84 ± 0.13 | 1.72 ± 3.22 | 1.88 ± 2.4 | 0.33 | 0.90 |
| ALT (IU/L) | 22.5 ± 8.5 | 21.4 ± 15.2 | 42.6 ± 33.2 | 0.51 | 0.02 |
| CK (IU/L) | NA | 69.5 ± 44.1 | 71.8 ± 50.8 | NA | 0.90 |
| D-dimer (μg/mL) | NA | 1.8 ± 2.0 (n=12) | 1.1 ± 0.4 | NA | 0.28 |
| Fbg (mg/dL) | NA | 389.0 ± 84.2 (n=7) | 480.4 ± 172.2 | NA | 0.22 |
| Hypertension | 2 (20.0%) | 6 (27.3%) | 3 (33.3%) | 0.70 | 1.00 |
| Diabetes mellitus | 0 (0%) | 5 (22.7%) | 3 (33.3%) | 0.17 | 0.66 |
| Dyslipidemia | 0 (0%) | 4 (18.2%) | 3 (33.3%) | 0.16 | 0.38 |
| Colonary heart disease | 0 (0%) | 2 (9.1%) | 0 (0%) | 1.00 | 1.00 |

Continuous variables were expressed as mean ± SD and tested by unpaired Student's *t*-test.
Categorical variables were expressed as n (%) and tested by Fisher's exact test.

**Table S4. EV proteins for discrimination between mild and severe COVID-19 patients.**

| GeneName | Cross-validation score | Sensitivity | Specificity | Accuracy | AUC |
|---|---|---|---|---|---|
| COPB2 | 1.00 | 1.00 | 1.00 | 1.00 | 1.00 |
| KRAS | 0.90 | 1.00 | 0.82 | 0.87 | 0.93 |
| PRKCB | 0.90 | 0.89 | 0.95 | 0.94 | 0.93 |
| RHOC | 0.90 | 0.78 | 1.00 | 0.94 | 0.96 |
| CD147 | 0.87 | 0.67 | 0.91 | 0.84 | 0.73 |
| CAPN2 | 0.87 | 0.89 | 0.77 | 0.81 | 0.84 |
| ECM1 | 0.87 | 0.67 | 0.95 | 0.87 | 0.82 |
| FGG | 0.87 | 0.67 | 1.00 | 0.90 | 0.87 |
| MFAP4 | 0.87 | 0.56 | 1.00 | 0.87 | 0.75 |
| ADI1 | 0.84 | 0.67 | 0.95 | 0.87 | 0.88 |
| AK1 | 0.84 | 0.89 | 0.91 | 0.90 | 0.95 |
| MGAT1 | 0.84 | 1.00 | 0.77 | 0.84 | 0.91 |
| CLDN3 | 0.84 | 0.89 | 0.82 | 0.84 | 0.86 |
| CRP | 0.84 | 0.78 | 0.77 | 0.77 | 0.82 |
| UQCRC2 | 0.84 | 0.78 | 0.82 | 0.81 | 0.77 |
| FGA | 0.84 | 0.67 | 0.95 | 0.87 | 0.88 |
| FGB | 0.84 | 0.67 | 1.00 | 0.90 | 0.84 |
| FGL1 | 0.84 | 0.78 | 0.82 | 0.81 | 0.85 |
| GPX1 | 0.84 | 0.67 | 0.91 | 0.84 | 0.81 |
| GSK3B | 0.84 | 0.44 | 1.00 | 0.84 | 0.73 |
| LBP | 0.84 | 0.78 | 0.86 | 0.84 | 0.82 |
| PDGFC | 0.84 | 0.89 | 0.77 | 0.81 | 0.86 |
| RAB13 | 0.84 | 0.78 | 0.86 | 0.84 | 0.85 |
| RAP1B | 0.84 | 0.67 | 1.00 | 0.90 | 0.91 |
| SLC6A4 | 0.84 | 0.89 | 0.82 | 0.84 | 0.90 |
| UBA7 | 0.84 | 0.78 | 0.86 | 0.84 | 0.83 |
| ORM1 | 0.81 | 0.89 | 0.59 | 0.68 | 0.80 |
| RNPEP | 0.81 | 0.56 | 0.95 | 0.84 | 0.68 |
| ANGPT1 | 0.81 | 0.78 | 0.86 | 0.84 | 0.88 |
| APOB | 0.81 | 0.78 | 0.86 | 0.84 | 0.79 |
| B4GALT1 | 0.81 | 0.67 | 0.95 | 0.87 | 0.79 |
| BHMT | 0.81 | 0.44 | 1.00 | 0.84 | 0.74 |
| CPN1 | 0.81 | 0.89 | 0.68 | 0.74 | 0.84 |
| GNAZ | 0.81 | 1.00 | 0.95 | 0.97 | 0.99 |
| ICAM2 | 0.81 | 1.00 | 0.59 | 0.71 | 0.83 |
| SELL | 0.81 | 0.67 | 0.91 | 0.84 | 0.74 |
| MAN1A1 | 0.81 | 0.78 | 0.82 | 0.81 | 0.85 |
| SERPINA5 | 0.81 | 0.89 | 0.82 | 0.84 | 0.81 |
| PACSIN2 | 0.81 | 0.89 | 0.86 | 0.87 | 0.90 |
| NCF1B | 0.81 | 0.89 | 0.55 | 0.65 | 0.73 |
| TMEM59 | 0.81 | 0.44 | 0.95 | 0.81 | 0.59 |
| YWHAB | 0.77 | 0.67 | 0.95 | 0.87 | 0.85 |
| ABAT | 0.77 | 0.33 | 1.00 | 0.81 | 0.52 |
| ADH1B | 0.77 | 0.67 | 0.95 | 0.87 | 0.79 |
| ASL | 0.77 | 0.67 | 0.91 | 0.84 | 0.80 |
| ASS1 | 0.77 | 0.78 | 0.86 | 0.84 | 0.79 |
| CDH2 | 0.77 | 0.56 | 0.95 | 0.84 | 0.69 |
| CAB39 | 0.77 | 0.89 | 0.91 | 0.90 | 0.94 |
| CPS1 | 0.77 | 0.67 | 0.82 | 0.77 | 0.74 |
| CD226 | 0.77 | 0.67 | 1.00 | 0.90 | 0.87 |
| COL6A3 | 0.77 | 0.67 | 0.86 | 0.81 | 0.82 |
| CUL4A | 0.77 | 0.78 | 0.64 | 0.68 | 0.75 |
| DSC1 | 0.77 | 0.44 | 0.95 | 0.81 | 0.58 |
| ENTPD5 | 0.77 | 1.00 | 0.64 | 0.74 | 0.86 |
| EIF4A1 | 0.77 | 0.67 | 0.86 | 0.81 | 0.80 |
| FN1 | 0.77 | 0.89 | 0.68 | 0.74 | 0.81 |
| PGC | 0.77 | 0.89 | 0.68 | 0.74 | 0.78 |
| RHEB | 0.77 | 1.00 | 0.59 | 0.71 | 0.84 |
| GNAI2 | 0.77 | 0.89 | 0.59 | 0.68 | 0.79 |
| GNB1 | 0.77 | 0.78 | 0.77 | 0.77 | 0.83 |
| GNA13 | 0.77 | 0.67 | 0.95 | 0.87 | 0.86 |
| ITGA2B | 0.77 | 0.67 | 0.91 | 0.84 | 0.86 |
| ITGB1 | 0.77 | 1.00 | 0.59 | 0.71 | 0.84 |
| ILK | 0.77 | 0.89 | 0.77 | 0.81 | 0.84 |
| F11R | 0.77 | 1.00 | 0.50 | 0.65 | 0.82 |
| LTA4H | 0.77 | 0.56 | 0.91 | 0.81 | 0.67 |
| LIMS1 | 0.77 | 0.89 | 0.77 | 0.81 | 0.83 |
| NAV2 | 0.77 | 0.56 | 0.86 | 0.77 | 0.73 |
| FAM129B | 0.77 | 0.78 | 0.86 | 0.84 | 0.84 |
| NNMT | 0.77 | 0.67 | 0.91 | 0.84 | 0.69 |
| NID1 | 0.77 | 0.89 | 0.55 | 0.65 | 0.76 |
| PPIA | 0.77 | 0.78 | 0.77 | 0.77 | 0.86 |
| PLA1A | 0.77 | 0.67 | 0.91 | 0.84 | 0.80 |
| PPBP | 0.77 | 0.67 | 0.77 | 0.74 | 0.69 |
| PECAM1 | 0.77 | 1.00 | 0.55 | 0.68 | 0.82 |
| GP1BB | 0.77 | 0.67 | 0.86 | 0.81 | 0.81 |
| PCSK9 | 0.77 | 1.00 | 0.82 | 0.87 | 0.91 |
| MENT | 0.77 | 0.44 | 0.95 | 0.81 | 0.71 |
| SERPINA10 | 0.77 | 0.67 | 0.82 | 0.77 | 0.74 |
| F2RL3 | 0.77 | 0.89 | 0.86 | 0.87 | 0.91 |
| LOX | 0.77 | 0.67 | 0.91 | 0.84 | 0.81 |
| SFTPB | 0.77 | 0.67 | 0.77 | 0.74 | 0.78 |
| RAB5B | 0.77 | 0.78 | 0.86 | 0.84 | 0.84 |
| RALB | 0.77 | 0.89 | 0.68 | 0.74 | 0.82 |
| REEP6 | 0.77 | 0.78 | 0.68 | 0.71 | 0.71 |
| RETN | 0.77 | 0.67 | 0.73 | 0.71 | 0.69 |
| AGXT | 0.77 | 0.22 | 1.00 | 0.77 | 0.48 |
| CCT2 | 0.77 | 0.89 | 0.64 | 0.71 | 0.81 |
| THBD | 0.77 | 0.56 | 0.91 | 0.81 | 0.74 |
| ISG15 | 0.77 | 0.33 | 1.00 | 0.81 | 0.67 |
| ZYX | 0.77 | 1.00 | 0.55 | 0.68 | 0.82 |

**Table S5. ExRNAs for discrimination between mild and severe COVID-19 patients.**

| Transcript | Cross-validation score | Sensitivity | Specificity | Accuracy | AUC |
|---|---|---|---|---|---|
| miR-122-5p | 0.84 | 0.56 | 1.00 | 0.87 | 0.81 |
| SNORD33 | 0.84 | 1.00 | 0.73 | 0.81 | 0.89 |
| AL732437.2 | 0.84 | 0.89 | 0.68 | 0.74 | 0.80 |
| RNU2-29P | 0.81 | 0.44 | 1.00 | 0.84 | 0.70 |
| CDKN2B-AS1 | 0.81 | 0.67 | 0.95 | 0.87 | 0.86 |
| AL365184.1 | 0.81 | 0.78 | 0.86 | 0.84 | 0.87 |
| AL365184.1 | 0.81 | 0.78 | 0.91 | 0.87 | 0.90 |
| AL365184.1 | 0.81 | 0.67 | 0.91 | 0.84 | 0.87 |
| AL365184.1 | 0.81 | 0.78 | 0.91 | 0.87 | 0.90 |
| AL365184.1 | 0.81 | 1.00 | 0.73 | 0.81 | 0.92 |
| let-7c-5p | 0.77 | 0.44 | 0.95 | 0.81 | 0.75 |
| miR-21-5p | 0.77 | 0.56 | 0.91 | 0.81 | 0.71 |
| miR-140-3p | 0.77 | 0.89 | 0.77 | 0.81 | 0.89 |
| C5orf66-AS2 | 0.77 | 0.78 | 0.91 | 0.87 | 0.89 |

**Table S6. Univariate Cox regrssion analysis of the selected factors and biomarkers for predicting COVID-19 severity in this patient cohort.**

| | Cut-off | HR | (95% CI) | P value |
|---|---|---|---|---|
| Age high | 56.5 | 28.1 | (3.4 - 231.9) | 0.0019 |
| Smoking index high | 87.5 | 3.1 | (0.8 - 11.6) | 0.092 |
| CRP high | 2.3 | 8.4 | (1 - 67.5) | 0.045 |
| ALT high | 21.5 | 3.8 | (0.8 - 18.5) | 0.095 |
| COPB2 low | $1.6 \times 10^{8}$ | NA | - | - |
| KRAS low | $4.8 \times 10^{7}$ | 189.8 | (0.4 - $9.0 \times 10^{4}$) | 0.095 |
| PRKCB low | $6.3 \times 10^{7}$ | 32.1 | (3.9 - 261.9) | 0.0012 |
| RHOC low | $1.4 \times 10^{7}$ | 23.6 | (4.7 - 118) | 0.00012 |
| CD147 high | $6.1 \times 10^{8}$ | 10.7 | (2.5 - 45.1) | 0.0013 |
| CAPN2 high | $7.2 \times 10^{5}$ | 15.5 | (1.9 - 125.9) | 0.010 |
| ECM1 high | $2.3 \times 10^{8}$ | 11.6 | (2.8 - 48.4) | 0.00079 |
| FGG high | $1.4 \times 10^{10}$ | 21.4 | (4.2 - 110.4) | 0.00025 |
| MFAP4 high | $4.6 \times 10^{8}$ | 12.7 | (3.3 - 48.6) | 0.00022 |
| miR-122-5p high | $1.0 \times 10^{5}$ | 10.5 | (2.7 - 40.4) | 0.00063 |
| SNORD33 high | 406.6 | 104.1 | (0.4 - $2.7 \times 10^{4}$) | 0.10 |
| AL732437.2 high | 8.8 | 9.9 | (1.2 - 79.9) | 0.031 |
| RNU2-29P high | 124.1 | 10.4 | (2.6 - 40.8) | 0.00081 |
| CDKN2B-AS1 high | 6.2 | 14.4 | (3.4 - 61.3) | 0.00031 |
| AL365184.1 high | 2.9 | 14.2 | (1.8 - 114.4) | 0.013 |

HR, hazard ratio; CI, confidence interval; NA, not applicable
